# A Crowdsourcing Approach to Develop Machine Learning Models to Quantify Radiographic Joint Damage in Rheumatoid Arthritis

**DOI:** 10.1101/2021.10.25.21265495

**Authors:** Dongmei Sun, Thanh M. Nguyen, Robert J. Allaway, Jelai Wang, Verena Chung, Thomas V Yu, Michael Mason, Isaac Dimitrovsky, Lars Ericson, Hongyang Li, Yuanfang Guan, Ariel Israel, Alex Olar, Balint Armin Pataki, RA2 DREAM Challenge Community, Gustavo Stolovitzky, Justin Guinney, Percio S. Gulko, Mason B. Frazier, James C. Costello, Jake Y. Chen, S. Louis Bridges

**Affiliations:** University of Alabama at Birmingham, Birmingham, AL, USA; Sage Bionetworks, Seattle, WA, USA; WRQ Research, 53 E 7 St #8, New York, NY, USA; Catskills Research, 1334 Hudson Pl, Davidson, NC, USA; Department of Computational Medicine and Bioinformatics, University of Michigan, 100 Washtenaw Avenue, Ann Arbor, MI, USA; Leumit Health Services, Tel-Aviv, Israel and Medil, solutions for digital medicine, Jerusalem, Israel; Eötvös Loránd University - Department of Complex Systems in Physics, Budapest, Hungary, Pázmány Péter sétány 1/A; IBM T J Watson Research Center, IBM, Yorktown Heights, NY, USA; Sema4, Stamford, CT, USA; Division of Rheumatology, Department of Medicine, Icahn School of Medicine at Mount Sinai, New York, NY, USA; Department of Pharmacology, University of Colorado Anschutz Medical Campus, Aurora, CO, USA; Division of Rheumatology, Department of Medicine, Hospital for Special Surgery, New York, NY, USA

## Abstract

To develop machine learning methods to quantify joint damage in patients with rheumatoid arthritis (RA), we developed the RA2 DREAM Challenge, a crowdsourced competition that utilized existing radiographic images and “gold-standard” scores on 674 sets of films from 562 patients. Training and leaderboard sets were provided to participants to develop methods to quantify joint space narrowing and erosions. In the final round, participants submitted containerized codes on a test set; algorithms were evaluated using weighted root mean square error (RMSE). In the leaderboard round, there were 173 submissions from 26 teams in 7 countries. Of the 13 submissions in the final round, four top-performing teams were identified. Robustness of results was assessed using Bayes factor and validated using an independent set of radiographs. The top-performing algorithms, which consisted of different styles of deep learning models, provided accurate and robust quantification of joint damage in RA. Ultimately, these methods lay the groundwork to accelerate research and help clinicians to optimize treatments to minimize joint damage.

## Introduction

Rheumatoid arthritis (RA) is a chronic inflammatory disease that affects ∼0.5-1.0% of populations worldwide, and more than 1.3 million people in the USA.^1^ The hallmark of RA is inflammation in the synovial lining of joints, leading to joint space narrowing, erosions in subchondral bone, and joint deformity.^2^ Joint damage in chronic RA is usually assessed over time through radiographs of the hands/wrists and feet, as these small joints are typically affected and have features that allow for differentiation from other forms of arthritis such as osteoarthritis. Quantitation of damage through imaging plays an important role in therapeutic decisions in individual patients, such as escalating therapy in patients with evidence of worsening erosions or joint space narrowing.

RA patients do not always develop joint damage, which reflects variability in the pathogenesis of the disease, clinical risk factors, and environmental exposures. In addition, some patients with minimal symptoms can slowly develop progressive radiographic damage over time, which may not be recognized by clinicians who see patients several times a year. Determining whether a patient is accruing joint damage and assessing the rate of progression are major challenges in treating patients with RA. While there are many advanced imaging techniques capable of quantifying RA damage (e.g. small coil MRI, ultrasound), these techniques are either expensive, unavailable to many patients, or operator dependent (subjective and dependent on the skill of the person acquiring the images). Thus, the most common approach to quantify RA-related bone and joint damage used in research studies and by many clinicians is radiographic images of hands/wrists and feet. The modified Sharp/van der Heijde (SvH) visual inspection scoring method is currently the gold standard for quantifying joint damage in RA.^3^ This method is validated for use in research studies, but cannot easily be applied to clinical practice, as the scoring is time-consuming, labor-intensive, and requires specialized training of readers. Thus, an automated way to quickly, accurately, and reproducibly assess the degree of joint damage would be highly valuable in clinical settings and would facilitate much larger research studies on factors associated with radiographic progress by enabling use of real-world data. One of the barriers to developing such methods is the lack of large sets of images with validated SvH scores for algorithm training and validation.

Algorithms based on deep learning have attained human-level or even better performance at image classification in recent Imagenet competitions.^4,5^ Neural networks or deep learning approaches have been applied to the diagnosis of pulmonary tuberculosis or pneumonia from chest radiographs.^6^ RA-related joint erosion and narrowing scoring represent a unique challenge due to the numerous joints involved (e.g. proximal interphalangeal, metacarpophalangeal, and many joints in the wrist) and the complex anatomy (e.g. overlapping carpal bones) (**Figure 1A**). Given advances in image analysis and machine learning, an increasing number of studies are trying to address the issue of automated scoring of radiographic images in RA.^7–9^ However, to date, no independently benchmarked, accessible and automated methods to quantify RA-associated joint damage in the hands/wrists and feet are in common use.

**Figure 1.**
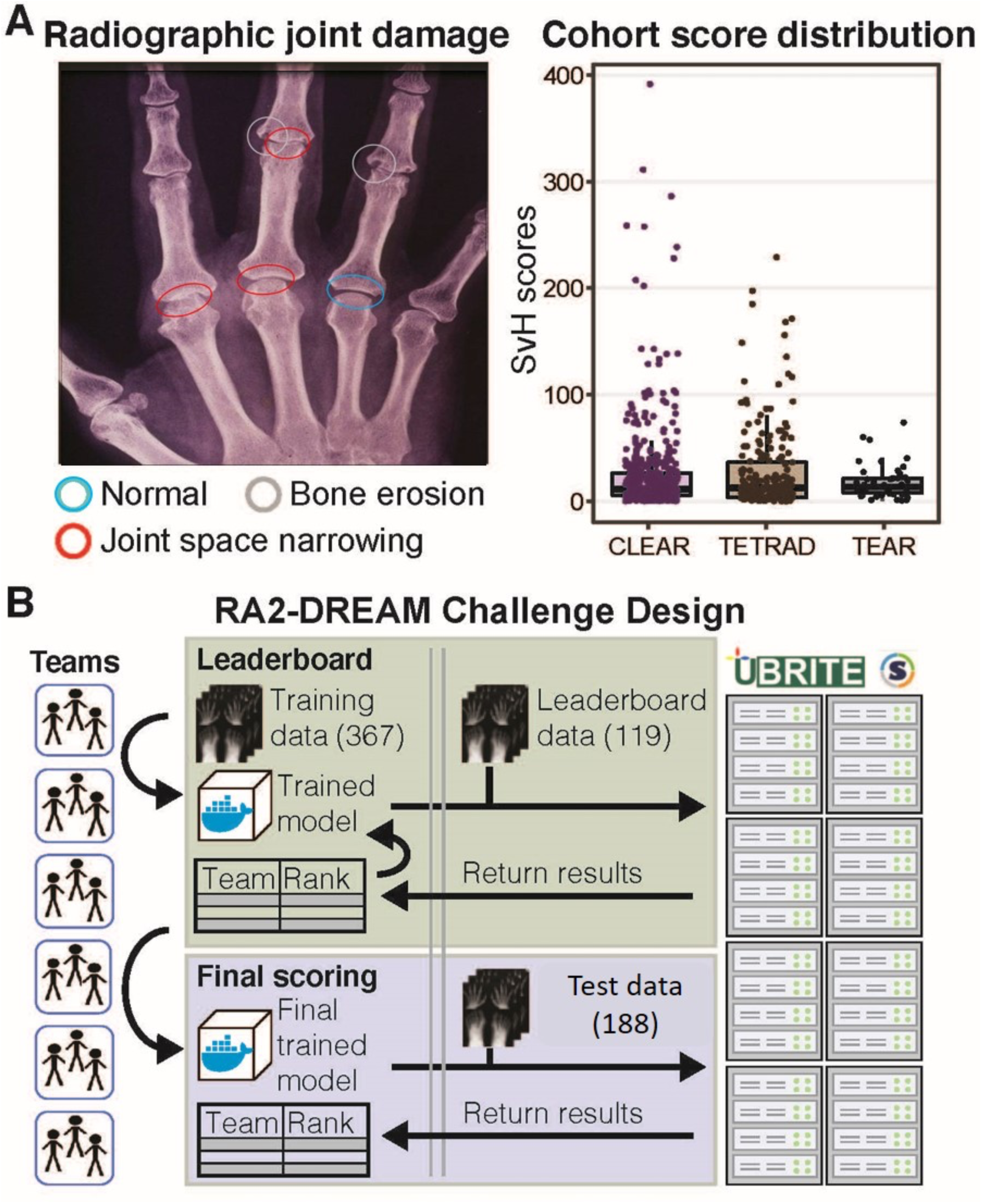
An overview of RA2 DREAM challenge. **A**. A representative radiograph of a RA patient showing normal joints (blue circled), joints with erosions (gray circled), and joint space narrowing (red circled). **B**. SvH score distribution of datasets used in the challenge and the post-challenge validation. Images with known SvH scores from CLEAR and TETRAD formed the training, leaderboard and final scoring round datasets. A subset of images from the TEAR trial was used for post-challenge method validation. **C**. A total of 367 sets of radiographic images and expert-curated SvH scores was provided to participants to train algorithms. Leaderboard data (n=119) and final scoring data (n=188) were used for performance evaluation in the leaderboard and final scoring rounds.

Here, we organized a crowdsourced, benchmarking effort to invite the worldwide community to develop robust methods for quantifying damage from RA radiographs. The RA2 DREAM Challenge, a community-based, collaborative competition engaged a diverse community of biologists, computer scientists, and physicians from all over the world. In this challenge, we provided radiographs generated from two NIH-funded studies with gold standard SvH scores, the Consortium for the Longitudinal Evaluation of African-Americans with Rheumatoid Arthritis (CLEAR)^10^ and the Treatment Efficacy and Toxicity in Rheumatoid Arthritis Database and Repository (TETRAD).^11^ Teams that participated in the RA2 DREAM Challenge were provided with access to high-resolution radiographic images of hands/wrists and feet. In the training set, accompanying SvH scores were also given, and teams developed computational methods to automatically and accurately score the overall RA-related damage, the degree of joint space narrowing, and the degree of erosions (Subchallenges 1, 2, and 3, respectively) (**Figure S1**). In the leaderboard phase, they were given a second independent set of films but scores were withheld; teams were given iterative feedback on the performance of their models. In the test phase, participants were given a third set of radiographic images (**Table 1**) and submitted two times but only one final model to be judged in the competition. ^12^

**Table 1.** Demographic characteristics of patients in training, leaderboard, and final scoring sets. Rheumatoid Factor and anti-CCP (anticyclic citrullinated peptide) antibody are two autoantibodies found in most RA patients. IQR, Interquartile range. SD, Standard deviation.

| Characteristics | Training Set<br>(N=367) | Leaderboard Set<br>(N=119) | Test Set<br>(N=188) |
| --- | --- | --- | --- |
| Overall Total Score<br>(median [IQR]) | 13.00<br>[5.00, 38.50] | 5.00<br>[2.00, 15.50] | 10.00<br>[5.00, 20.00] |
| Joint Erosion Score<br>(median [IQR]) | 5.00<br>[2.00, 17.00] | 2.00<br>[1.00, 5.00] | 5.00<br>[2.00, 9.00] |
| Joint Narrowing Score<br>(median [IQR]) | 7.00<br>[2.00, 22.50] | 2.00<br>[0.00, 11.00] | 4.00<br>[0.75, 11.25] |
| Sex (%), Female | 307 (83.7) | 108 (90.8) | 158 (84.0) |
| Race (%) |  |  |  |
| Black or African-American | 242 (65.9) | 88 (73.9) | 156 (83.0) |
| White | 106 (28.9) | 26 (21.8) | 21 (11.2) |
| Others | 19 (5.2) | 5 (4.2) | 11 (5.9) |
| Age (Years, mean (SD)) | 54.87 (13.24) | 51.36 (13.12) | 53.45 (13.46) |
| Age having RA<br>(Years, mean (SD)) | 46.27 (13.31) | 45.91 (13.94) | 48.41 (13.07) |
| Disease Duration<br>(Months, mean (SD)) | 103.18 (113.06) | 65.39 (74.25) | 60.45 (93.88) |
| Rheumatoid Factor (%) |  |  |  |
| Positive | 273 (74.4) | 89 (74.8) | 126 (67.0) |
| Negative | 90 (24.5) | 29 (24.4) | 60 (31.9) |
| NA | 4 (1.1) | 1 (0.8) | 2 (1.1) |
| anti-CCP Antibody (%) |  |  |  |
| Positive | 250 (68.1) | 76 (63.9) | 112 (59.6) |
| Negative | 94 (25.6) | 39 (32.8) | 68 (36.2) |
| NA | 23 (6.3) | 4 (3.4) | 8 (4.3) |

There were a total of 13 final valid submissions, each of which was evaluated against manually curated scores using a patient-weighted root mean square error (RMSE) metric. We included a baseline model for comparison (see Methods). A robustness analysis was performed to compare the performance against the baseline model and top performers. The algorithms of four teams were identified as top performers for the three subchallenges. Top-performing algorithms and others were then validated using an additional independent set of images with known SvH scores from the Treatment of Early Aggressive Rheumatoid Arthritis (TEAR) trial.^12^ As we show, these methods acheived performance that is comparable to human scorers. This supports automated scoring of RA radiographs as a feasible and promising approach that may be used both in research and clinical medicine in the near future.

## Results

### The top-performing teams for overall damage (SC1), JSN (SC2), and erosion scores (SC3)

The top-performing algorithms were determined based on their weighted RMSE. The stability or robustness of the models was evaluated by bootstrap analysis. Bayes’ factor was used to determine if subsequently ranked teams were tied with the top performing team (Bayes factor <= 3); no ties were observed (**Figure 2**). Top performing teams across all subchallenges had better performance than the baseline model. Team Shirin, Hongyang Li/Yuanfang Guan, csabaibio and Team Gold Therapy’s developed the top-performing models to predict overall damage (**Figure 2A**), joint space narrowing (**Figure 2B**), and erosion (**Figure 2C**). Results were further confirmed by calculating pairwise p-values (**Figure S2 A-C**). Detailed writeups of individual methods are supplied in the **Supplemental Methods** and **Table S1**.

**Figure 2.**
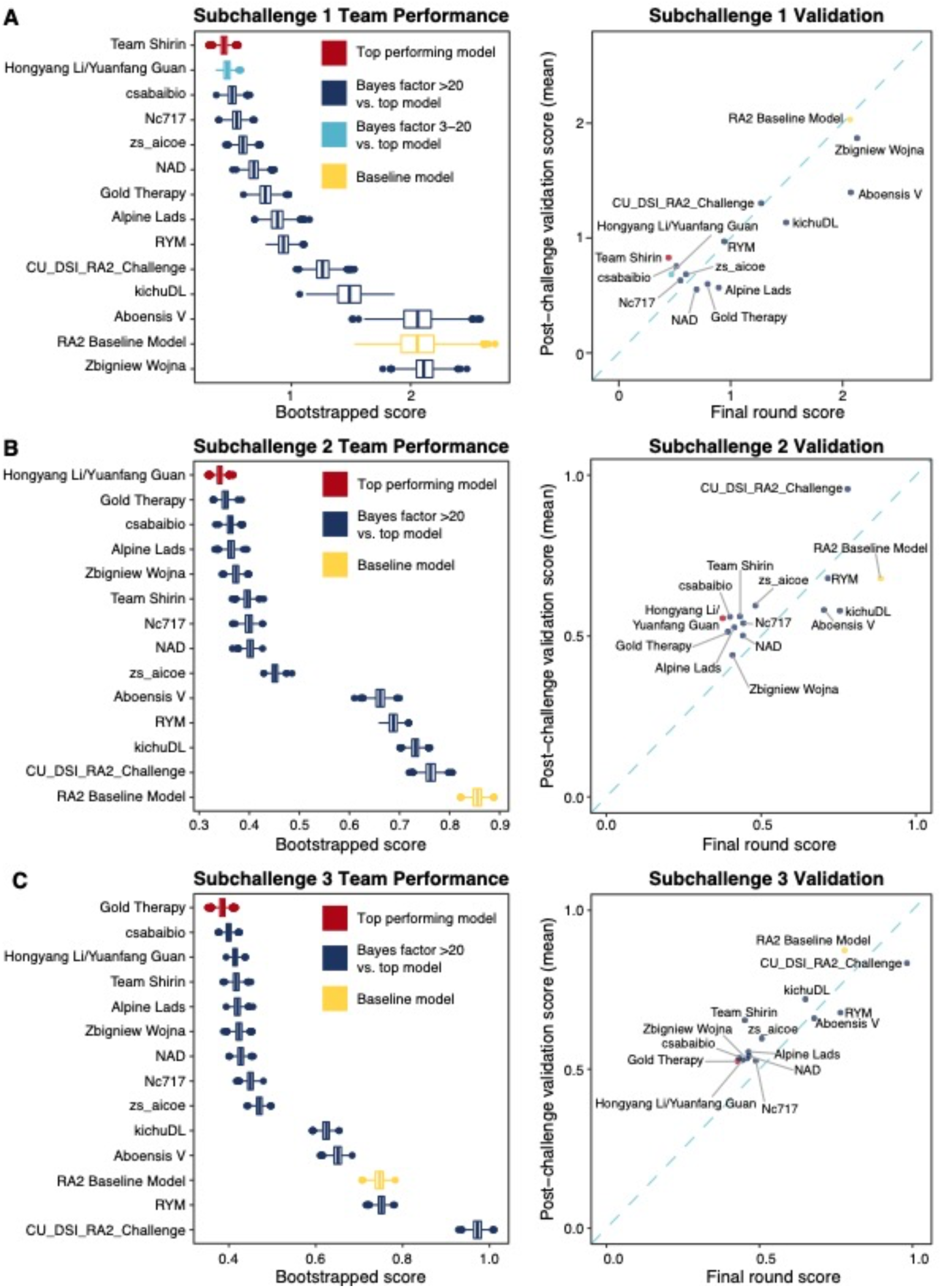
Evaluation and validation of challenge results. (**A, B, C, left**) The performance of each team in SC1, SC2, SC3. All predictions were bootstrapped to generate a distribution of scores and to calculate Bayes factors between the top performing model (red) and all other models. Any prediction with a Bayes factor of 3 or less was considered “tied” with the top model; no predictions were tied for top place in any of the three subchallenges. The baseline model provided by the organizers was used for reference (yellow). (**A, B, C, right**) The models were run on an independent, post-challenge validation dataset of 50 image sets and scored against two sets of gold standard measurements from the validation dataset, using the SC1, SC2, and SC3 metrics, respectively (**A, B, C**); the dotplots show the weighted RMSE performance in the final scoring round (x-axis) for each model compared to the mean weighted RMSE between two sets of gold standard validation measurements from the post-challenge validation set (y-axis). Algorithms below the dashed blue line performed better in the final scoring round, while those above the blue line performed better on the validation dataset.

### The top-performing teams results are rigorous and reproducible

Submitted algorithms may depend on random initialization^24^ or contain other stochastic components. To assess the reproducibility and stability of each method, we re-ran each algorithm on the final scoring round dataset. We found that all the top performing teams (and indeed, most of the algorithms) produced identical (Spearman = 1) or nearly identical (Spearman > 0.99) predictions between the final scoring round results and the rerun scores (**Figure S3**). Thus, all submitted models were highly reproducible with stable predictions.

We then evaluated generalizability of methods to other datasets using an independent, post-challenge validation dataset from the TEAR Trial. The coefficient of variation for SvH overall, JSN, and erosion between two readers in the TEAR Trial were 0.74 (p=0.15), 0.62 (p=0.18) and 0.73 (p=0.03). The TEAR study contained slightly lower quality radiographs than those from the CLEAR and TETRAD studies, suggesting that the post-challenge validation task was likely more difficult than the final round. We calculated the weighted-RMSE and compared the algorithms’ performances in to the final scoring round (**Figure 2**).We also computed a concordance index between the final scoring round scores and the post-challenge validation dataset scores. A concordance index of 1 indicates perfectly concordant rankings, while a concordance index of 0 indicates perfectly discordant (i.e. opposite) ranking. The concordance indices were 0.714 for SC1, 0.78 for SC2, and 0.824 for SC3. This demonstrates that the methods predicted consistently between the final scoring dataset and the post-challenge validation dataset, suggesting that these algorithms can be readily applied to new, independent radiographs.

There were some inconsistencies between top-performing algorithms in the final scoring dataset compared to the post-challenge validation dataset. Team NAD in SC1 and Zbigniew Wojna in SC2 showed better performance than others in the post-challenge validation, which may be due to better performance in certain scenarios. For example, the NAD and Zbigniew Wojna models may perform better than others in images with different types of artifacts, images of different resolution/quality, or datasets with differences in which joints had more damage (e.g. wrists vs PIP joints). This also suggests that combinations of algorithms may be more robust than any individual model alone.

### Ensemble models are better than each individual team’s model

As described in the Methods, we generated ensembled predictions from the models submitted in the final scoring round. We ensembled the top two performers, then added subsequently ranked teams over all models (**Figure 3**). We then performed the same bootstrapping analysis to assess robustness of the ensemble models and calculated Bayes factor compared to the top performing team.

**Figure 3.**
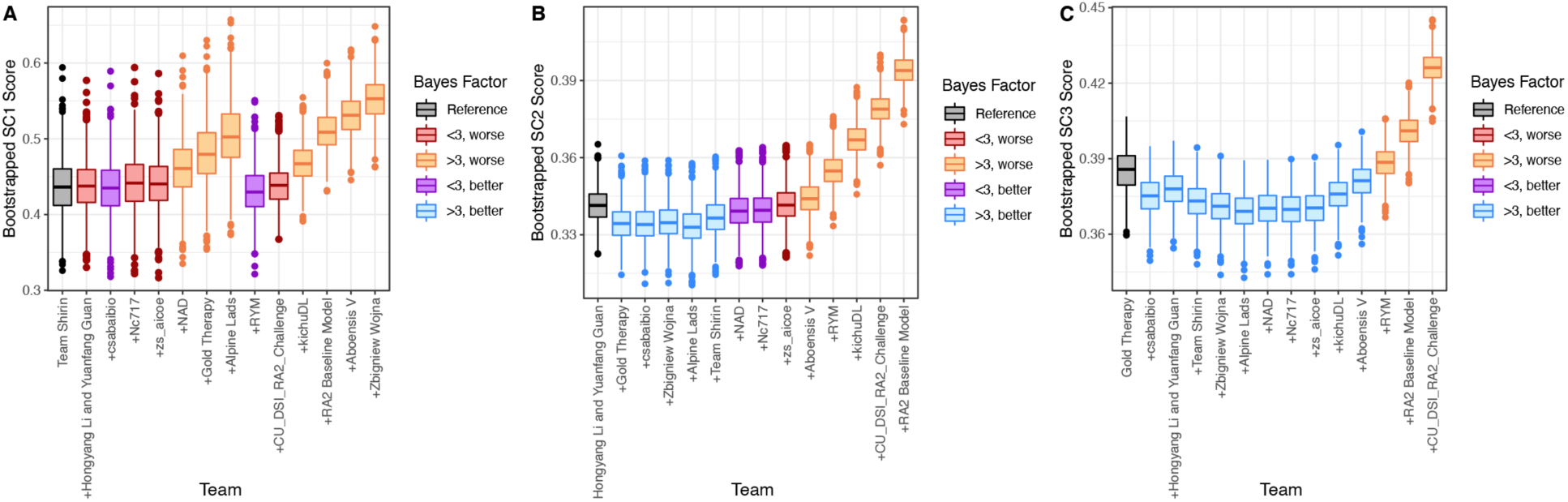
Ensembled models improve performance (weighted RMSE). A series of simple ensemble models were created by combining the top two models, the top three, and so on until all models were combined. For each ensemble model, the predictions were averaged (mean) and scored with the SC1, SC2, or SC3 metrics. A bootstrap/Bayes factor analysis was used to determine differences in performance between the top-performing (individual) model and the ensemble models. We labeled ensemble models in blue/purple when the absolute score was “better” than the top model, and orange/red when the absolute score was “worse” than the top model. The purple or red indicates models that have a Bayes < 3 compared to the top model and were thus considered tied, while the blue or orange indicates Bayes > 3 and therefore models that are substantially better or worse, respectively than the top-performing model alone. **A**. For SC1, all ensemble models perform equivalently (Bayes < 3, re/purple) or worse than the top model alone (Bayes > 3, orange). **B**. For SC2, the first 5 ensemble models perform better than the top model alone (Bayes > 3, blue). **C**. For SC3, the majority of the ensemble models are better than the top model alone (Bayes > 3, blue).

For SC1, the ensembled models were not substantially better than the top-performing method; that is, no ensembled models had a bootstrapped lower weighted RMSE with a Bayes factor > 3 relative to the top-peforming model (**Figure 3A**). Interestingly, including predictions from team RYM substantially improved performance, even though it was not one of the top-ranked models in SC1. After evaluating the distribution of the predictions (**Figure S4A**), we determined that the ensemble model that included the top-performing algorithm and the predictions from RYM (and the 7 algorithms ranked between them) systematically predicted greater damage for all patients. When combined with the better-performing models, this systematic alteration brought the distribution of the predictions closer to that of the gold standard (**Figure S4B**).

For SC2 and SC3, we observed that averaging the predictions from several of the top-ranked models (up to 6 and 10, respectively) increased the predictive power of the algorithms substantially (Bayes factor > 3). We also evaluated the ensemble predictions using an unweighted metric (Spearman correlation, **Figure S5**). Ensembling several top models yielded similar improvements in performance in all three subchallenges.

### Impact of case-by-case technical decisions on team performance

To gain insights into the submitted methods and to aid future algorithm development, we systematically summarized the top-performing teams’ methods by conducting a post-challenge survey and evaluating the submitted method write-ups (**Table 2**). We received responses from 11 of the 13 teams who submitted algorithms in the final scoring round. Among these 11, two teams did not apply image segmentation before scoring the joint damage. In general, teams who applied image segmentation achieved better performance for all three subchallenges (**Figure S6A**), though the difference between using and not using segmentation is not statistically significant (KW test p-value > 0.1). For the methods that did perform segmentation, no specific algorithm or publicly available segmentation models were associated with method performance (**Figure S6B**).

**Table 2.**
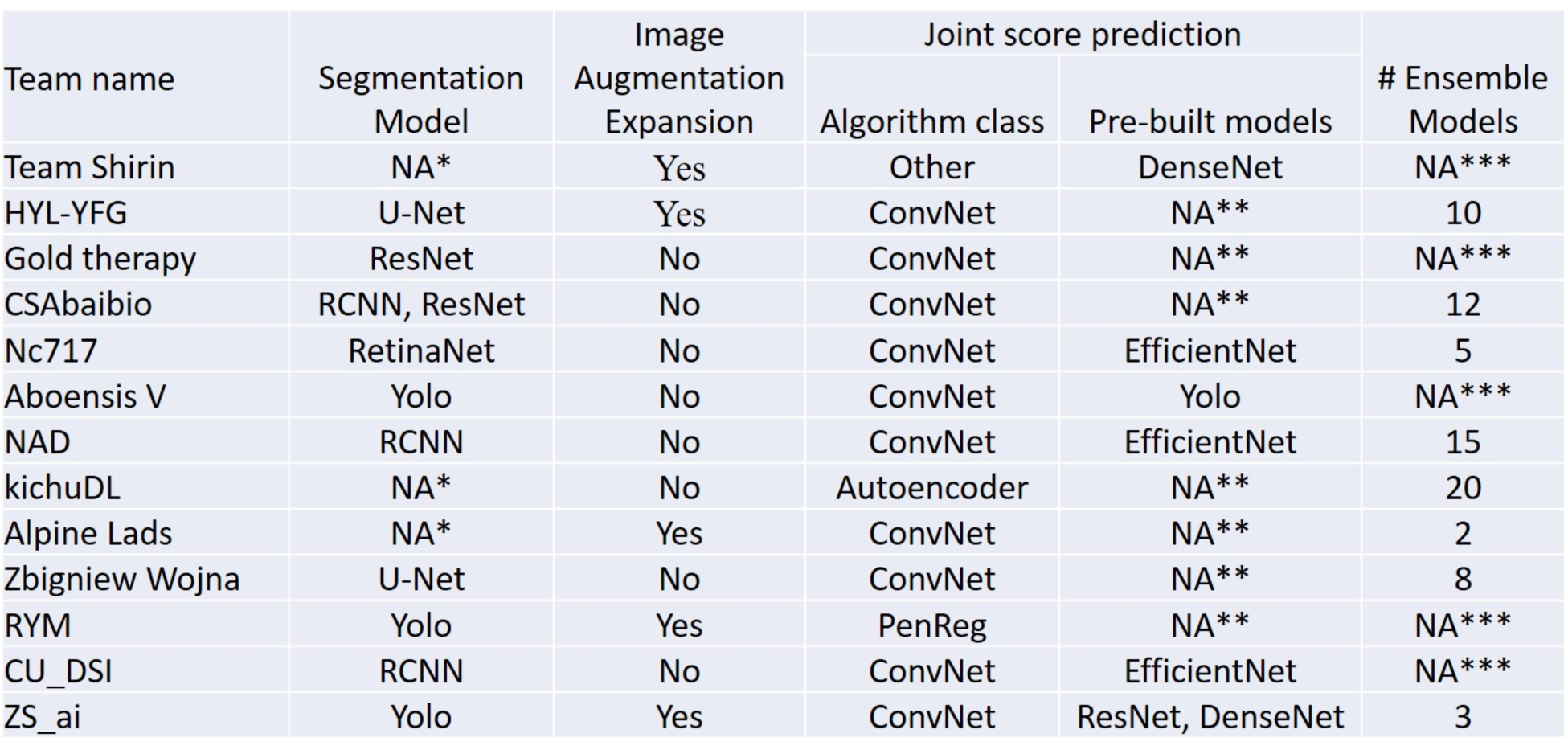
Summary of machine learning methods used by the final-round teams. NA*: the team did not apply segmentation. NA**: the team did not apply any prebuilt model. NA***: the team did not use ensemble model. See **Table S2** for more details.

Nine of the 11 teams applied deep learning-based approaches. The average scores among algorithms that used deep learning were more favorable than those not using deep learning methods, but this difference failed to reach statistical significance (KW test p-value > 0.1) (**Figure S6C**). We did however find that applying ensemble approaches improved team performances in SC2 and SC3 (KW test p-value=0.06; **Figure S6D**).

## Discussion

In the RA2 DREAM Challenge, we leveraged the international community of computer science and engineering experts to develop methods to automatically score overall RA severity, joint space narrowing, and erosions on radiographic images of the hands/wrists and feet. There is complexity and variability inherent in the joint images on RA radiographs. For example, many joints of the wrist are overlapping on images, and are not aligned in the same plane as the radiation beam, leading to subjective interpretation. This likely explains our finding that scoring of the MCP and PIP joints in the hands and the joints of the forefoot were more accurately scored than those in the wrist.

In addition, we found that scores for joint space narrowing were more accurate than those for erosions. After our challenge was closed, we identified outlier participant-produced scores compared to expert-curated SvH scores by comparing joint space narrowing and erosion scores of individual joints on the same radiograph (see **Supplemental Methods** and **Figure S7** and **Table S3**). We found 201 outliers of 7,896 individual joint narrowing scores (2.5%) and 462 outliers of 8,272 (5.6%) individual joint erosion scores. One of our investigators (MBF, a board certified musculoskeletal radiologist) re-scored the confirmed outliers. He made corrections to the SvH scores of 97 (1.23%) of joint space narrowing scores and 192 (2.32%) of erosion scores in the final scoring dataset. This low number of “corrections” needed in the expert curated scores show that there were few “mistakes” made by the trained scorers. In the analysis of TEAR validation data the coefficients of variation between the two expert readers were 0.74, 0.62 and 0.73, for overall, joint space narrowing, and erosion, showing more variability in the erosion scores than in the joint space narrowing scores. This also suggests that joint space narrowing is easier to assess than erosion and that the overall score is largely determined by the least predictive component. Difficulties in scoring of erosions may be due in part to the difficulty in visualizing the bony cortex precisely in all joints on a given radiographic image. Future challenges may help to overcome this difficulty by developing automated optimization of images through digital manipulation of brightness, contrast, and allowing for targeted high-resolution magnification.

The vast majority of the RA2 Challenge participants used deep learning-based methods, reflecting the general trend of deep learning application in image analysis. This adoption rate is likely due to the availability of freely accessible pre-trained models (such as DenseNet,^25^ ResNet,^26^ and U-Net^27^), which were used by all of the top-performing teams, as well as the flexible and extendable architecture of these methods. The flexibility of these methods may lead to the dependence of the final performance on other technical decisions beyond using a deep learning framework. Importantly, building ensemble models substantially improves the predictive performance and is likely to be included in highly successful future approaches to analysis of radiographic images.

Our results provide optimism for future automated scoring, but larger-scale follow-up studies are needed. In order for algorithms to become truly robust, many thousands of images should be used for training and models should be continuously benchmarked on new data. The successful implementation of algorithmic scoring could make millions of stored images without JSN and erosion scores available for research on factors associated with radiographic damage in RA. While the compilation of the CLEAR, TETRAD, and TEAR images is a valuable first step, future directions should include digitally acquired annotated images from large-scale observational studies, clinical trials, and EHR datasets. Significantly expanding our capability to study joint damage at a significantly lower cost and with greater reproducibility creates new possibilities for understanding the pathogenesis of RA and the development of new treatments to prevent joint damage.

Lessons learned in this DREAM Challenge may contribute to quantification of radiographic damage in other types of arthritis in which erosions have different characteristics than RA. This might include such as pencil-in-cup deformities in psoriatic arthritis or “punched-out” erosions with overhanging edges in gout. In addition, automated approaches may ultimately help clinicians make therapeutic decisions, such as changing treatment regimens to avoid additional joint damage.

This RA2 DREAM Challenge showed that international collaboration and award-incentivized crowd-sourcing can lead to robust and reproducible algorithms to score radiographic damage in RA. Furthermore, it exemplifies how community-based team science may engage both domain experts and machine learning and artificial intellegence experts in the future.

## Methods

### Challenge data

Two NIH-funded clinical studies, CLEAR^10^ and TETRAD^11^, led by investigators at the University of Alabama at Birmingham (UAB), provided radiographic images and standard SvH scores for the RA2 DREAM Challenge.

We used a total of 674 sets of images from 562 patients with corresponding SvH scores. There were 455 sets of images/SvH scores from 408 CLEAR participants and 209 sets from 154 TETRAD participants (**Figure 1A**). A total of 47 CLEAR and 55 TETRAD participants had sets of images at two different time points, typically about 1 year apart. Each set comprised four images: one of each foot and one of each hand/wrist, in a postero-anterior direction. SvH scores^13^ were previously generated by experienced readers trained in this method, as has been reported.^10, 11^ The SvH scoring system assesses 15 areas from each hand/wrist and 6 areas from each foot to assess JSN; and 16 joint areas from each hand/wrist and two sides of 6 joints from each foot to evaluate erosions. Each area is assigned a score from 0 to 4 for narrowing and 0 to 5 for erosion, leading to a total score of 448 (the overall total narrowing score is 168, and the overall erosion score is 280).

Importantly, we included a substantial number of joints without RA-associated joint damage, which was important for training algorithms to recognize joints with no JSN or erosions. Approximately ∼34% of the films from CLEAR participants were scored 0^14^ because many had early RA and not all patient develop damage. There were fewer TETRAD participants with no damage because that study focused on the treatment to refractory RA, leading to more damage. In addition TETRAD participants had longer disease duration, on average, than CLEAR participants.

### Challenge Procedures

Each of the three subchallenges were evaluated separately using the expert-curated SvH overall (SC1), JSN (SC2), and erosion (SC3) scores as the gold standard. In the training round, participanting teams were provided with a set of images and accompanying SvH scores for each joint to develop their algorithms. In the leaderboard round, only images were provided, and teams refined their algorithms through sequential submissions and scoring feedback after assessments by the RA2 DREAM Challenge organizers. In the final scoring round, only images were provided and one final containerized submission was submitted for evaluation and performance ranking of teams. Each of the sets (training, leaderboard, and final scoring) were independent of each other, with no sets of images used in more than one round.

The socio-demographic and clinical characteristics of the participants whose radiographs were used are shown in **Table 1**. The distributions of SvH scores from the three studies utilized in this challenge (CLEAR, TETRAD, TEAR) are shown in **Figure 1B**. In the leaderboard round, each team was required to submit a containerized version of their method for evaluation and received weighted RMSE scores as feedback. Public leaderboards were updated immediately after the performance of each algorithm was assessed. In the leaderboard round, each team was allowed up to three submissions per week over 9 weeks, totalling 27 potential submissions (**Figure S1**). This allowed teams to iteratively evaluate the performance of their algorithms and modify accordingly. In the final round, teams were allowed to submit up to two containerized models (with the last submission considered to be a team’s final submission) and were required to include a written description of their final method. Each submission was evaluated to assess performance and to rank each model according to how well it performed relatively to the expert-curated scores (**Figure S1** and **1C**).

All teams submitted their models via the Synapse collaborative science platform in the form of containerized code (a Docker container). The containers were required to follow a prescribed format to enable automated execution in the University of Alabama at Birmingham (UAB) Cheaha supercomputer. Additionally, the models were required to run without a network connection and had to define an ENTRYPOINT to run the algorithm. Participants were allowed to train their models locally and submit a trained model or explore the training data locally and submit an untrained model to be trained at run-time. Models submitted to Synapse were automatically transferred to the UAB Cheaha supercomputer using the CWL Synapse Workflow Hook, converted to Singularity containers, and executed on the challenge data to produce prediction files (**Figure 1C**).

### Challenge Evaluation

For each model, the DREAM Challenge organizing team produced the overall (SC1), JSN (SC2), and erosion (SC3) SvH scores. We assessed the performance of each team’s model by comparing the scores derived from their methods (*y*_*i*_ in the equation below) to the ground truth SvH scores (*s*_*i*_ in the equation below) using a patient-weighted root mean square error (RMSE). First, we assigned a weight (*w*_*i*_ in the equation below) to each patient according to their log transformed overall SvH score. Specifically, weight 1, 2, 2.14, 3.86, 8, 16, 32 and 64 were assigned to an overall SvH scores or score ranges of 0, 1, 2-3, 4-7, 8-20, 21-55, 56-148, and >148, respectively (**Table S4**). We used the weights to generate equal-weighted sets of data into the training (n=367, radiographic images and scores), leaderboard (n=119, radiographic images only), and final scoring (n=188, radiographic images only) datasets. We then calculated a patient-weighted RMSE using the RMSE of log-transformed team scores and expert-curated SvH scores, multiplied by the patient weights (*w*_*i*_), and then calculated the average RMSE across the entire cohort:

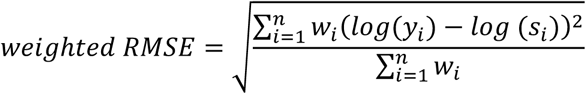

### Baseline model

A baseline model (https://www.synapse.org/#!Synapse:syn21570587) was developed by the RA2 DREAM Challenge organizers for comparison to the models submitted by the participants. We created the baseline model by training a 10-layer deep learning model without segmentation of the images to classify the damage (0-4 for narrowing and 0-5 for erosion). The JSN and erosion scores were summed to generate SC1 scores. The layer architecture followed the example described in Chapter 5 of *Deep Learning with Python*.^15^

### Robustness analysis and tied methods assessment

The robustness of each submitted algorithm (and the baseline model) was determined using Bayes factor analysis. This analysis identifies which methods that are better than, tied with, or worse than a reference method (e.g. the top-performing submission, or a baseline submission) considering the performance on random sampling of the data. In both the final scoring and post-challenge validation rounds (see following section), we performed this analysis using the top-performing model (to identify ties), as well as the baseline model (to identify which models performed ‘better’ than the baseline). Briefly, we applied bootstrapped sampling^16^ to create 1000 subsets from the original data set. In each subset, we reran the two models for comparison to obtain scores for SC1/SC2/SC3. We then used these scores to calculate Bayes factors for each subchallenge using the “computeBayesFactor” function of R package “challengescoring” (https://github.com/sage-bionetworks/challengescoring). Smaller Bayes factors indicate more similar performance. As has been done with previous challenges, Bayes factor greater than 3 was considered significantly different.^17–22^ False discovery rate (FDR) corrected pairwise Student’s t-test p-values for each team relative to the top-scoring team in each subchallenge were also calculated as a secondary comparison of performance.

In addition, we interrogated the stability of predictions when different random computational initialization was applied on the same dataset. The re-run was executed after announcing the final round results and did not impact the team ranking. We independently re-ran each model submitted in the final scoring round once and computed the Spearman correlation between the final scoring round and post-challenge validation round scores.

### Post-challenge Validation

To validate the performance of all algorithms, we selected 50 independent, high quality image sets with accompanying SvH scores from the TEAR Trial.^12^ In the TEAR Trial, there were two sets of SvH scores for each set of images, each generated by one of two independent readers. We ran all containerized models submitted on the 50 independent sets of quantile-normalized images and evaluated their performance compared to the mean SvH measurements from each of the two readers. We also computed the Spearman correlation between the final scoring and post-challenge validation dataset to assess the reproducibility of the top-performing algorithms. The concordance index between the final scoring and post-challenge validation rounds was calculated using the concordance_index function in the Python “lifelines” library.

### Post-challenge survey

To summarize the array of approaches used in the RA2 DREAM Challenge, we sent a post-challenge survey to all participating teams (see questions in **Supplemental Methods** and **Table S2**) and analyzed the results of the survey by grouping teams’ scores based on their responses to each question. We then applied the Kruskal-Wallis (KW) test ^23^ among the groups to evaluate whether the different methodological choices were significantly associated with performance.

### Ensemble modeling

As with previous DREAM Challenges, we explored the ‘wisdom of the crowds’ phenomenon by performing ensembling experiments with the models submitted for the final scoring round.^17–22^ We generated multiple ensembled predictions by calculating the mean prediction for each joint across multiple algorithms. This produced ensemble predictions that, for each subchallenge, aggregated the top performer alone, the top two, the top three, and so on. We then scored these ensemble predictions using the previously described Bayes factor robustness analysis.

## Data Availability

All radiogrpahic images used in this study are available upon reasonable request to the authors. Method write-ups and Docker containers for all submitted methods are available from the Challenge workspace (www.doi.org/10.7303/syn20545111).

https://www.synapse.org/#!Synapse:syn20545111/wiki/594083

## Data and code availability

The use of the Challenge data (images and SvH scores) was approved by the UAB Institutional Review Board for Human Use. The scoring code Docker container is available from the Synapse Docker repository (<u>docker.synapse.org/syn20545112/scoring_harness:latest</u>). Method write-ups and Docker containers for all submitted methods are available from the Challenge workspace (www.doi.org/10.7303/syn20545111). Please refer to Synapse documentation (https://help.synapse.org/docs/Synapse-Docker-Registry.2011037752.html) for guidance on downloading Docker containers from the Synapse Docker Registry.

## RA2 DREAM Challenge Community Authors

**Team Zbigniew Wojna:** Zbigniew Wojna [1], Anna Krason [2]; **Team RYM:** YanMing Tan [3], RaphaelHaoChong Quek [3]; **Team Nc717:** Neelambuj Chaturvedi [4]; **Team AlpineLads:** Michael Stadler [5], Chenfu Shi [5]; **Team kichuDL:** Krishnakumar Vaithinathan [6], Julian Benadit P [7]; **Team NAD:** Duc Tran [8], Tin Nguyen [8]**; Team Gold Therapy:** Ariel Israel [9] [10]; **Team Hongyang Li and Yuanfang Guan:** Hongyang Li [11], Yuanfang Guan [11]; **Team Shirin:** Isaac Dimitrovsky [12], Lars Ericson [13]; **Team csabaibio:** Bálint Ármin Pataki [14], Alex Olar [14]; **Team Aboensis V:** Alexander Biehl [15][*], Mehrad Mahmoudian [15][16][*], Sami Pietilä [15][*], Tomi Suomi [15], Mikko S Venäläinen [15][*], Laura L Elo[15][17]; **Team zs_aicoe**: Chenguang Xue [18], Akshat Shreemali [18], Srinivas Chilukuri [18]; **Team CD_DSI_RA2_Challenge**: Khanh-Tung Nguyen-Ba[19], Jay Ji-Hyung Ryu[19], Rui Bai[20], Yilin Wu[19], Yingnan Wu[20], Xiaofu He[20][21]

[1] Tensorflight, Inc.350 5th Avenue, Suite 4215, New York, NY 10118, USA

[2] University College London, Gower St, London WC1E 6BT

[3] NUS Department of Statistics and Data Science, 21 Lower Kent Ridge Rd, National University of Singapore, Singapore 119077

[4] ZS Associates, Safina Towers, Bengaluru, India

[5] Centre for Genetics and Genomics Versus Arthritis. Division of Musculoskeletal and Dermatological Sciences, School of Biological Sciences, Faculty of Biology, Medicine and Health, The University of Manchester, UK

[6] Department of Computer Engineering, Karaikal Polytechnic College, Varichikudy, Karaikal-609609, Puducherry, India

[7] Department of Computer Science and Engineering, School of Engineering and Technology, CHRIST(Deemed to be University), Kengeri campus, Kanmanike, Bangalore-560074, India

[8] Department of Computer Science and Engineering, University of Nevada, Reno, NV 89557, USA

[9] Leumit Health Services, Tel-Aviv, Israel

[10] Medil, solutions for digital medicine, Jerusalem, Israel

[11] Department of Computational Medicine and Bioinformatics, University of Michigan, 100 Washtenaw Avenue, Ann Arbor, MI, USA

[12] WRQ Research, 53 E 7 St #8, New York NY 10003

[13] Catskills Research, 1334 Hudson Pl, Davidson NC 28036

[14] Eötvös Loránd University - Department of Complex Systems in Physics, Budapest, Hungary, Pázmány Péter sétány 1/A

[15] Turku Bioscience Centre, University of Turku and Åbo Akademi University, Turku, Finland

[16] Department of Future Technologies, University of Turku, Turku, Finland

[17] Institute of Biomedicine, University of Turku, Turku, Finland

[18] ZS Associates Inc, 1560 Sherman Ave, Evanston, IL, 60201

[19] Department of Statistics, Columbia University, Room 1005 SSW, MC 4690, 1255 Amsterdam Avenue, New York, NY 10027

[20] Data Science Institute, Columbia University, Northwest Corner, 550 W 120th St, #1401, New York, NY 10027

[21] Department of Psychiatry, Columbia University, 1051 Riverside Drive, New York, NY 10032

[*] equal contributors to Team Aboensis V

## Acknowledgments

We would like to acknowledge funding support from Bristol Myers Squibb (BMS) and the CTSA Program National Center for Data to Health (CD2H) is supported by the National Center for Advancing Translational Sciences (NCATS) at the National Institutes of Health (Grant U24TR002306).

