## Supplemental Methods for "A Crowdsourcing Approach to Develop Machine Learning Models to Quantify Radiographic Joint Damage in Rheumatoid Arthritis"

**Computational identification of gold standard outliers**

SvH scoring of radiographs by visual inspection is a labor-intensive process and can be error-prone. One strategy for identifying these errors is to compare a collection of algorithmically-predicted SvH measurements to visual inspection measurements from the same radiograph. For a given joint score, if the predicted scores from many good models (from challenge participants) are relatively similar and the manual SvH score is an outliner to the predicted scores, then this joint score is more likely an error. We used this concept to identify potential outliers or measurement errors in the test dataset (**Figure S7**). We used a binning strategy (**Figure S7 A-C**) to enable the calculation of a false discovery rate (FDR)-corrected empirical p-value for each gold standard (SvH) measurement in the test dataset. We binned each individual measurement (total overall, joint space narrowing and joint erosion scores) for each joint based on the mean.

After the challenge was completed and for each subchallenge, we took the mean of the top 8 predictions for a given overall score or joint score and assigned each joint to a bin based on this mean predicted score using the “cutr” R package smart_cut function to generate equal sized-bins. We calculated an empirical p-value for each gold standard SvH measurement relative to the other values in the same bin. We then false discovery rate (FDR)-corrected the p-values. Potential outlier gold standard measurements were defined as those that had an FDR-corrected p-value < 0.1. A board-certified musculoskeletal radiologist reviewed these flagged joints to determine whether the original measurements were inaccurate. If the review indicated that the measurement was incorrect, we re-scored these joints for joint space narrowing and erosion (**Figure S7 D-F**, pink bars and **Table S4**). We did not identify any putative errors in the gold-standard scores used for SC1 (overall damage) (**Figure 7D, Table S4**). For joint space narrowing (SC2) and erosion (SC3), we determined that 201/7896 (2.5%) and 462/8272 (5.6%) scores, respectively, were possible errors in the gold standard dataset (**Figure S7 E-F**). After the outliers were identified, a visual examination of the sets of radiographs with the highest discordance between submitted scores and SvH scores was performed to adjudicate whether the algorithm-generated score or the expert-curated score was more accurate. The adjudication was performed by a board-certified musculoskeletal radiologist (MBF), who reviewed the 663 (201 joint space narrowing, 462 erosion) discrepant measurements to determine whether the original measurements were indeed inaccurate, and if they were, to revise these joint scores for joint space narrowing and erosion to more accurately reflect ‘truth’. We created an in-house Matlab visualization package to overlay the scores on the images and increase the speed of the evaluation manual revision process. Using this approach, we were able to visualize and evaluate the accuracy of the gold-standard dataset. This re-evaluation of the radiographs resulted in correction of 97/201 (48%) of discrepant joint space narrowing and 192/462 (42%) of discrepant erosion scores (**Table S4**). This represented a correction of only 97/7896 (1.23%) joint space narrowing and 192/8272 (2.32%) of erosion scores overall. The corrected gold-standard was used for all post-challenge analysis.

**Evaluating the impact of final-round teams' technical decisions on the performance**

To better understand the submitted methods to aid future algorithm development and software development, we systematically summarized the winning teams' methods through collection of answers to following questions (Table 2, Table S2 and Figure S6):

- Which pre-built model the teams used in the segmentation? The options include: NA (not applying segmentation), self-built, or the name and citation of other pre-built software libraries.
- Whether the team enriched the 'rare class', i.e., severe patients, in the image augmentation step? Image augmentation is the common processing step in image recognition, when synthetic images are created to increase the number of training datapoints.
- Which class of algorithms the teams applied in predicting the joint damage score? The options include: Linear regression, Penalized regression (LASSO, Ridge, etc), deep learning, decision tree - random forest, and other.
- Which pre-train models and software library the teams used to predict the join damage score.
- How many sub-models the teams built when using the ensemble approach? Briefly, the ensemble approach builds many prediction models for the same task; the final prediction is the aggregation of all sub-models' result.

**Finalist team methods (write ups)**

All finalist teams applied deep-learning-based methods in both segmentation (when applicable) and joint score prediction steps. The teams manually located and marked the joint regions to train the segmentation model in the segmentation step. The deep-learning-based segmentation model may directly learn to draw the joint bounding boxes or the pre-defined points marking the joint. Pre-trained deep-learning-based models, including DenseNet [2], U-Net [3], ResNet [5], Efficient-Net [7], YOLO v3 [8], and Faster RCNN [10], were intensively retrained and repurposed for the DREAM Challenge dataset. Besides, the following practice improved the model performance

- *XGBoost* [9] is a regularizing gradient boosting software library that implements deep learning algorithms' core optimizer. Compared to other gradient boosting implementations, XGBoost is explicitly for distributed computing, which means multiple computers can solve the optimization together. This implementation has the advantage in cloud computing and supercomputing platforms, such as in Cheaha.
- *Hyperparameter selection*. The performance of the deep-learning-based model depends on the hyperparameters (such as initial weights), which have to be set before computing the models. This step requires manual decisions from each team. In practice, each team prepared some hyperparameter sets. They randomly split the training data into two parts. One part was used to compute the model with the selected hyperparameter set; and the other was used to test and compare the performance of the hyperparameter sets.
- Ensemble model. Some teams trained multiple independent joint score prediction models, then combined these models' results to determine the final joint score. There were two approaches to combine the models' results. The averaging/voting approach considered all models equal to each other. Then, the final score was determined by averaging all results. The sequential approach has one classification model and some regression models. When combining, the classification model determines whether the joints have damage; here, '0' means that the joints have no damage, and the final joint scores are 0. Otherwise, the regression models, which predict the damaged joint scores from 1 to 5, determine the final joint score.
- *VGG* [12] is a software library to create and train a new deep-learning model. The VGG-based deep learning network has 16 or 19 layers. As mentioned above, this library is used when the team decides not to retrain and repurpose the pre-trained models.

- *Autoencoder* is a deep-learning architecture that matches two homogeneous data sets, such as translation between two languages or synthesizes the data. The autoencoder synthesizes 'fake' data such that the classification algorithms may not differentiate between the 'fake' and the 'original data. The deep autoencoder includes an input 'original' data layer, an output 'fake' data layer, and many layers between. The center layer, which is also called the encoded layer, has the smallest size. The result at this layer is the most compacted the highest-quality representation of the original data yet; therefore, the encoded layer is used as features for later machine learning models.

***Shirin Team***

We first trained models to rotate/flip all images into the same 90 degree orientation, allowing for differently oriented input images and also effectively doubling the training set size (left and right images).

Joint centers were then manually marked on the training set, and this was used to train models to locate hand and foot joints on radiographs. For the finger and toe joints each joint was marked separately. The 12 wrist joints were grouped into 4 center points representing 3 joints each.

Joint sub-images were cut out using the above joint location models by taking a square image at the predicted joint center that was a fixed percentage of the original images (12.5% for fingers and toes, 15% for thumb/big toe, 25% for wrist). These joint sub-images were used to train models to predict erosion and joint narrowing scores. Groups of joints were merged to increase training data per model; for example, a single model was trained to predict all finger joint erosion scores, resulting in several thousand training examples for that model.

In addition to more commonly used data augmentation transforms (brightness, zoom, rotation, etc), perspective warps were used to simulate joints held at different angles to the X-ray plane. Gaussian blurring [1] was also found to improve performance, particularly on erosion prediction. Joints with more severe damage were duplicated to help with imbalance in the training set.

Models for orientation and joint center location used a pretrained Resnet34 architecture with 256x256 image size. Models for joint damage prediction used a pretrained Densenet201 architecture [2], also with image size 256x256 (note this was the size of the joint sub-images cut from the original images as described).

Five-fold cross-validation was used to manually select the best hyperparameters for joint damage prediction models. Single models were then trained using the best hyperparameter settings and all training data, and used for test set prediction. The final SvH score was added from all joint scores.

***Hongyang Li and Yuanfang Guan Team***

All images were uniformly scaled to 1024x1024-pixel size. Each joint in the image was manually marked by a 30x30 pixel bounding box. Image background noise filtering was applied. Two types of deep-learning models were trained. First, for image segmentation, there was a model trained for each joint. Each model was retrained using the U-Net (kernel size 7) pre-trained model and the joint bounding-box sub-images. Each U-Net [3] used three input channels: the original, gray-scaled image, the post-quantile-normalization image, and the post-noise-filtering image. Second, for joint score prediction, new deep-learning-based models were trained using seven input channels. The input channels include: (1) the overall original black-and-white image, (2) the overall image after quantile normalization, (3) the overall image after rescaling to the range between 0 and 255, (4) the overall image after filtering out background and quantile normalization, (5) the overall image after Gaussian normalization, (6) the bounding-box joint image after rescaling to the range between 0 and 255 only based on image patches, and (7) the bounding-box joint image after Gaussian normalization. Four joint score prediction models were built using deep learning to predict: (1) joint space narrowing in hand, (2) joint space narrowing in foot, (3) joint erosion in hand, and (4) joint erosion in foot. To predict the final patient SvH score, the joint scores were used as features to train an ensemble random forest model. The random forest consisted of 500 random decision trees.

In the testing phase, all the above input channels were computed. The U-Net retrained segmentation model automatically located the 30x30 pixel bounding box for each joint. Each joint score was predicted using the bounding boxes and image channels above as features. These predicted joint scores were fed toward the random forest model to predict the patients' SvH scores.

***Csabaibio Team***

From the challenge training image, horizontal flipping, 10-degree random rotating, and randomly changing 5% of the brightness and contrast were applied to increase the training set size improve the model robustness. In each image, the joints were manually marked by rectangle bounding boxes. The deep-learning-based and pre-trained Mask-RCNN [4] and ResNet-50 [5] were combined and trained using the bounding-box sub-images for image segmentation. All sub-images were rescaled to the size of 256x266 pixels. For joint score prediction, each joint score was the average of 12 independent deep-learning-based models. These 12 models were retrained from the following pre-trained models (citation): ResNet-34 (6 models), ResNet-50 (4 models), and Vgg16 (2 models). The training hyperparameters were manually selected by 5-fold cross-validation.

In the testing phase, the segmentation model automatically located the bounding boxes for the joints in each image. The bounding-box sub-images were rescaled to the size of 256x266 pixels. Then, the ensemble prediction model used the bounding-box images to make the joint score prediction. The patient's final SvH score was added from all joint scores.

***Nc717 Team***

The joints in the training images were manually marked by rectangle bounding boxes and labeled. The bounding boxes were used to train two segmentation models: one for hand images and the other for foot images. In segmentation, the RetinaNet/ResNet [5, 6] pre-trained model was retrained using the joint images. In each joint score prediction, five models were retrained from the pre-trained Efficient-Net model [7]. For each joint score, the final score was averaged from the results of these 5 models.

In the testing phase, the segmentation model automatically located the bounding boxes for the joints in each image. Then, the ensemble prediction model used the bounding-box images to make the joint score prediction. The patient's final SvH score was added from all joint scores.

***Zs_aicoe Team***

The joints in the training images were manually marked by rectangle bounding boxes and labeled using YOLO v3 [8] software. The images and bounding boxes were used to retrain YOLO v3 [8] model for segmentation. Segmentation retraining continued until the segmentation 'Intersection over union' metric reached 0.9 for hand images and 0.88 for foot images. Joint sub-images inside the bounding boxes were randomly rotated and shifted by a small amount; this created more images to train the joint score model later. Different pre-trained models and ensemble approached were applied to train models that predict different types of joint scores. –

- For erosion joint scores, the pre-trained ResNet-50 model was retrained into three models. The ResNet-50 [5] classification-based model considered each joint score is a binary class: 0 (no damage) versus other (with damage, erosion score from 1 to 5). The ResNet-50 regression-based model considered each joint score as a continuous number. So does the ResNet-XGBoost [9] regression-based model technique (citation).

To combine three model results into one erosion joint score, first, the classification-based result was examined. If the classification-based model predicted class 0, then the final erosion joint score is 0. Otherwise, the average result of the two regression-based models was used to determine the discrete damage score.

- For narrowing scores, only the ResNet-XGBoost [9] regression-based model was retrained and predict the joint score. No ensemble approached was applied.

In the testing phase, the segmentation models automatically located the bounding boxes for the joints in each image. The ensembled ResNet-50 and ResNet-XGBoost models predicted the erosion scores; the ResNet-XGBoost model predicted the narrowing scores. The patient's final SvH score was added from all joint scores.

***NAD Team***

From the challenge training image, horizontal flipping, small-degree random rotating, small shifting was applied to increase the training set size improve the model robustness. Thirty-four bounding boxes were manually marked in each patient's image, including 12 feet-joint boxes (6 left, 6 right), 20 finger-joint boxes (10 left, 10 right), and 2 boxes that covered the patient wrist joints (1 left, 1 right). The images and bounding boxes were used to retrain the Faster RCNN [10] pre-trained model for segmentation. In predicting the joint scores, 15 independent models were retrained from the EfficientNet [7] pre-trained model. These 15 models were created by 15 rounds of random splitting in the training set. For each joint score, the final score was determined by voting the output of these 15 models.

In the testing phase, the segmentation models automatically located the bounding boxes for the joints in each image. The ensembled 15 models predicted the joint scores using the sub-images inside the bounding boxes. The patient's final SvH score was added from all joint scores.

***Gold Therapy Team***

From the challenge training image, horizontal flipping, small-degree random rotating, and randomly changing a small amount of the brightness and contrast were applied to increase the training set size improve the model robustness. Each image was marked by a pre-determined set of anchor points, as showed in https://www.synapse.org/#!Synapse:syn21499370/wiki/604451. These anchor points located the joints' locations in the image and were the target of the pre-trained ResNet segmentation model. Using the pre-determined anchor points as the reference, after ResNet-based [5] segmentation model found these corresponding points in the patient images, the model could uniformly draw the bounding boxes, scale, and rotate the bounding-box sub-images to the same size and orientation. For each joint damage score, the regression-based pre-trained ResNet-35 [5] and RestNet-50 models [5] were retrained and enhanced by XGBoost [9]. Models' hyperparameters were determined empirically using hyperopt [11].

In the testing phase, the segmentation models automatically located the anchor points in each image. The joint bounding boxes were automatically extracted, scaled, and rotated from the detected anchor points. The ensembled ResNet-35 and RestNet-50 [5] models predicted the joint scores by averaging the output of these models. The patient's final SvH score was added from all joint scores.

***Alpine Lads Team***

Each image was marked by a pre-determined set of landmark points, as showed in https://www.synapse.org/#!Synapse:syn21610007/wiki/604496. These landmarks located the joints' locations in the image and were the target of the VGG-based model [12]. The landmarks define bounding-box regions that surround the joints in the images. These bounding-box sub-images were rescaled to the size of 224x224 pixels and fed to the extended-VGG model for each joint score prediction. Before training extended-VGG model [12], the sub-images were randomly zoomed by a small amount to increase the number of training samples. In predicting the joint damage score, the extended-VGG model consisted of two parts. The first part was VGG-based binary classification, which determined whether the joint had damage (0 for no damage, 1 for joint score from 1 to 5). Images that received the classification of '0' had the joint score of 0. Meanwhile, images that received the classification of '1' were further fed to the VGG-based regression model, where the model result determined the joint damage score.

In the testing phase, the segmentation models automatically located the landmarks in each image. The 224x224-pixel joint bounding boxes were automatically extracted from the detected landmarks. The two-stage VGG-extended model predicted the joint scores by averaging the output of these models. The patient's final SvH score was added from all joint scores.

***RYM Team***

All training images were cropped to remove unnecessary background. For the hands, the bottom 1/7 of each image were removed. This mostly removed the beginnings of the ulna and radius bones and part of the wrist. For the feet, the bottom ¼ of each image was removed. This did not remove any of the joints involved in scoring as the toes are found nearer the top of the feet. After cropping, the training images were scaled to the size of 1500 x 1200 pixels. A U-Net [3] model was trained to remove the nonuniform background intensity, which was considered noise prior to segmentation. Then, the joints in the image were manually marked by rectangle bounding boxes. Two pre-trained YOLOv3 [8] segmentation models were retrained for segmenting these joint bounding boxes; one segmented the hand images, and the other segmented the foot images. To predict each joint score, a VGG-based model was created and trained using the corresponding joint sub-images.

In the testing phase, the U-Net model above filtered out the image noise. Then, the YOLOv3 segmentation models automatically located the joint bounding boxes in each image. The VGG-based models predicted the joint scores. The patient's final SvH score was added from all joint scores.

***CU_DSI_RA2_Challenge Team***

The joints in the training images were manually marked by rectangle bounding boxes and labeled. For image segmentation, the deep-learning-based and pre-trained FasterCNN [10] was retrained using the bounding-box sub-images. The bounding boxes were scaled to the size of 224x224 pixels. For predicting each joint score, the pre-trained EfficientNet [7] was retrained using the 224x224-pixel sub-images. Before training, the sub-image data were augmented by randomly cropping, horizontal flipping, rotating, and distorting by a small amount.

In the testing phase, the FasterCNN segmentation model automatically located the 224x224-pixel joint bounding boxes in each image. The EfficientNet models predicted the joint scores. The patient's final SvH score was added from all joint scores.

***KichuDL Team***

Contrast Limited AHE [13] was applied in all images to remove the background noise. Then, all images were scaled to the size of 128x128 pixels. Then, a deep autoencoder was trained from these 128x128-pixel images. In this autoencoder, the input had the size 128x128, which corresponded to the scaled input image. The output layer had the same size as the input layer, which corresponded to the synthetic image. The middle layer, which was the smallest one, had the size of 32x32. After passing through the autoencoder, the results at the 32x32 middle layer were used to training other deep-learning models to predict the joint scores. Here, for each joint score, 20 independent models were trained and ensembled.

In the testing phase, first, Contrast Limited AHE filtered the image background noise. Second, the image was fed into the autoencoder from the input to the middle layer. Third, the output at the middle layer was fed into the deep learning models to predict the joint scores. The patient's final SvH score was added from all joint scores.

***Aboensis V Team***

All images were scaled to the size of 800x800 pixels. The joints in the training images were manually marked by rectangle bounding boxes and labeled using YOLO v3 [8] software. The images and bounding boxes were used to retrain YOLO v3 model for segmentation. Then, to predict each joint score, a pre-trained YOLO v3 model was retrained using the bounding-box sub-images.

In the testing phase, the YOLO v3 segmentation model automatically located the joint bounding boxes in each image. The YOLO v3 prediction models predicted the joint scores. The patient's final SvH score was added from all joint scores.

***Zbigniew Wojna Team***

The joints in the training images were manually marked by rectangle bounding boxes and labeled. The images and bounding boxes were used to retrain UNet [3] model for segmentation. Then, to predict each joint score, 8 pre-trained UNet models were retrained using the bounding-box sub-images. The final joint score was the average of these 8 models.

In the testing phase, the UNet segmentation model automatically located the joint bounding boxes in each image. Eight UNet prediction models predicted and ensembled the joint scores. The patient's final SvH score was added from all joint scores.
