## Supplemental Figures for "A Crowdsourcing Approach to Develop Machine Learning Models to Quantify Radiographic Joint Damage in Rheumatoid Arthritis"

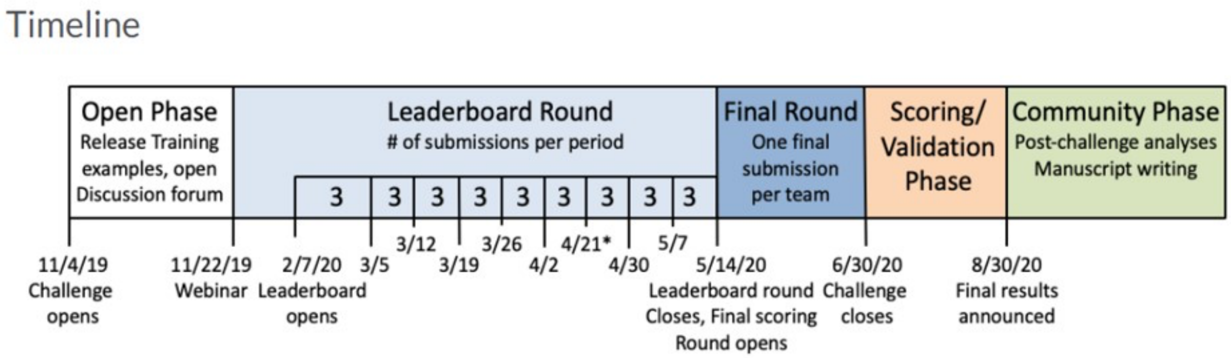


**Figure S1. RA2 DREAM Challenge timeline.**

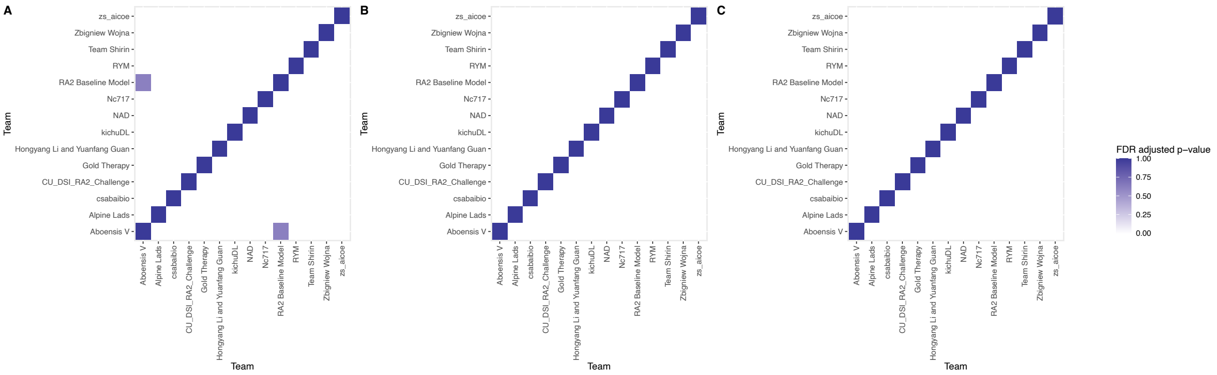


**Figure S2. Pairwise p-values between all final round predictions.** This analysis was performed for **A.** SC1, **B.** SC2, and **C.** SC3, and indicates that there is no statistical similarity between any challenge submissions (FDR-adjusted $p < 0.05$).

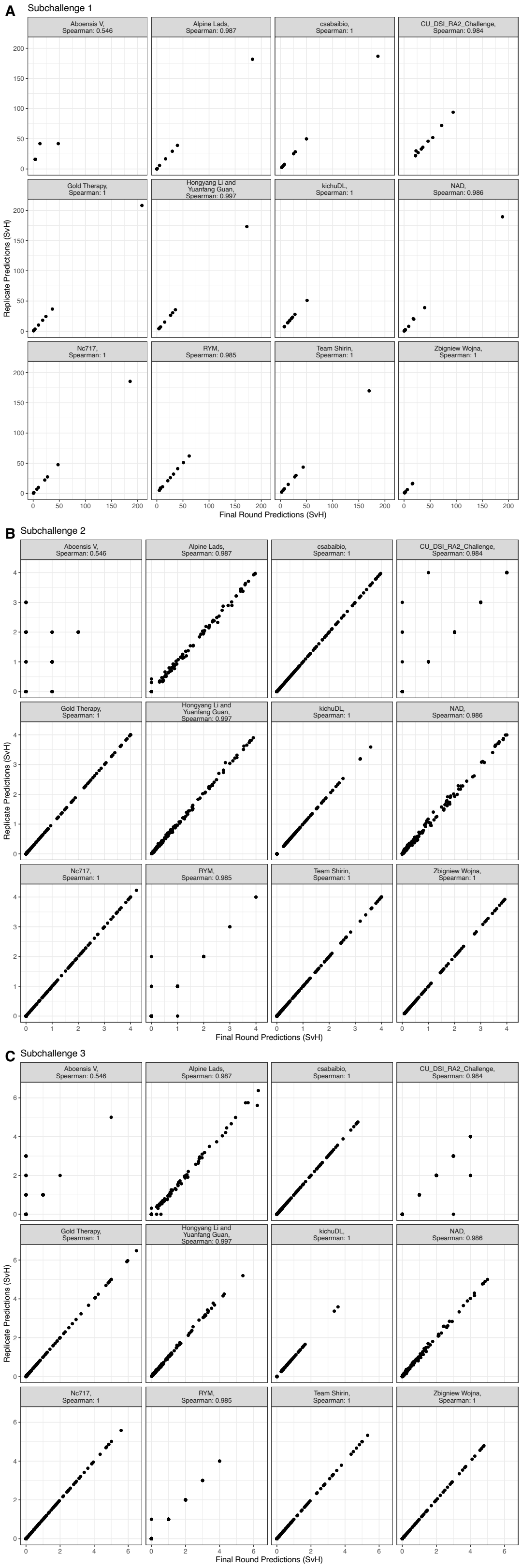


**Figure S3. Reproducibility of submitted algorithms.** Spearman correlation between the final round score and the post-challenge rerun of the same method; both use the same test data; **A.** SC1, **B.** SC2, and **C.** SC3.

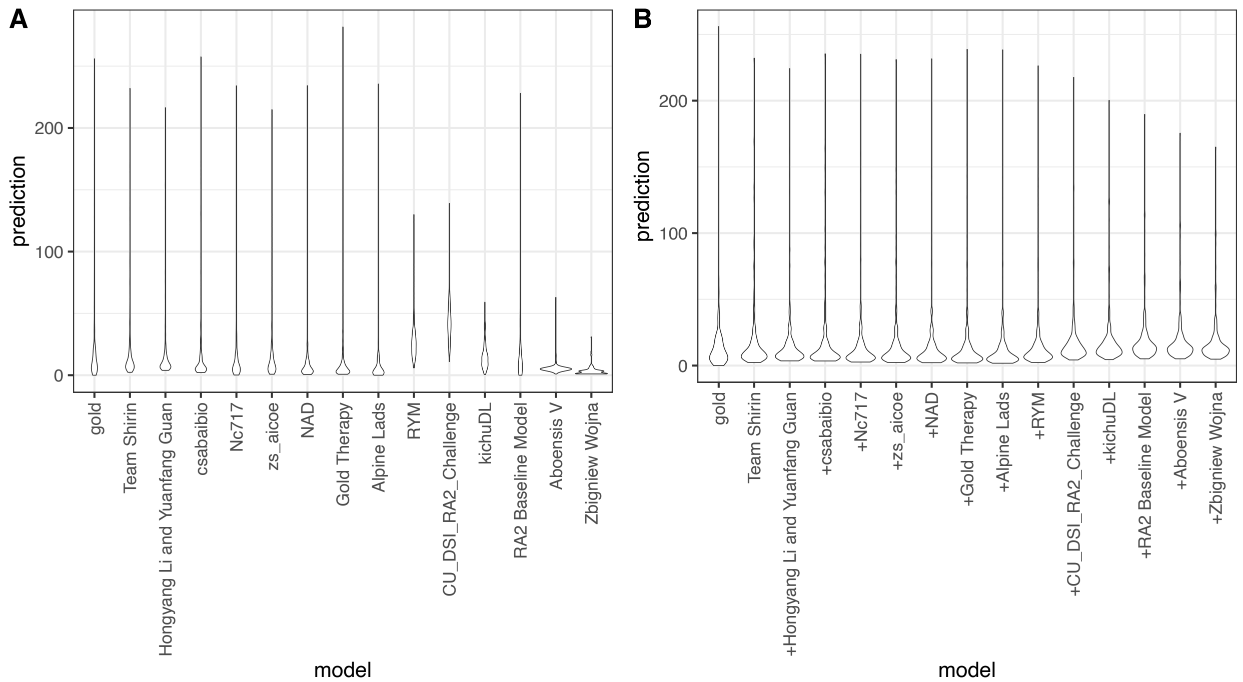


**Figure S4. Distribution of individual and ensembeled predictions**. **A.** The distribution of the submitted predictions (SC1,2,3 measurements) for each team. Compared to better-ranked predictions, the RYM model predicted greater damage for all patients. **B.** The distribution of the ensembeled predictions (SC1,2,3 measurements). When ensembled, the addition of the RYM model made the distribution of the predictions closer to that of the gold standard.

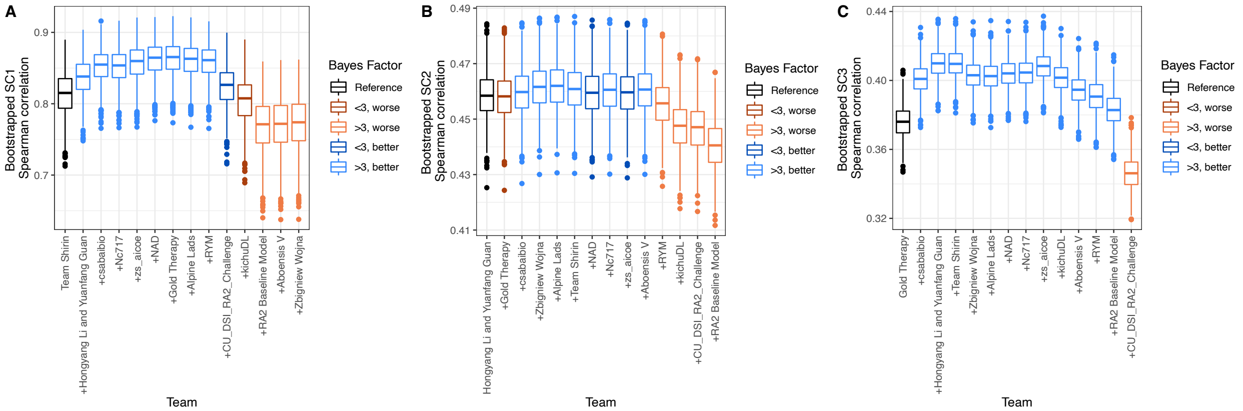


**Figure S5. Ensembled models improve performance (Spearman correlation).** The analysis described for **Figure 3** was repeated using Spearman correlation as the performance metric instead of weighted RMSE. Using this metric, we observed improved performance (Bayes < 3, light blue) for many of the ensembled models in **A.** SC1, **B.** SC2, and **C.** SC3.

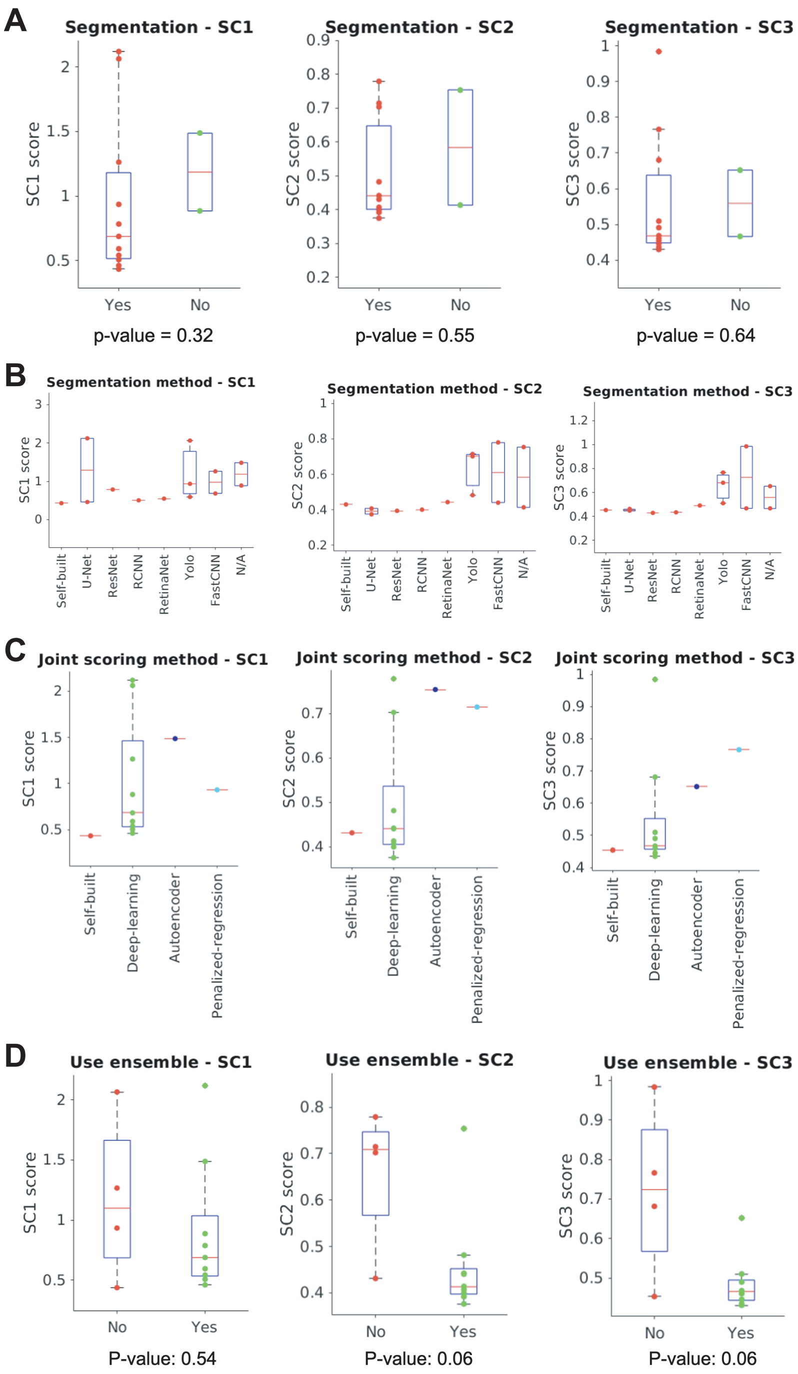


**Figure S6. Summarizarion of approaches and methods used by participated teams in three subchallenges**. **A.** Impact of segmentation on the subchallenge scores. Teams using vs. not using segmentation methods were compared. **B.** Impact of segmentation algorithms on the subchallenge scores. Participants self-built segmentation method, U-net, ResNet, RCNN, RetinaNet, Yolo, FastCNN and without any segmentation method were compared. **C.** Impact of scoring algorithm on the subchallenge scores. Participants self-built algorithm, Deep-learning, Autoencoder and Penalized-regression were compared. **D.** Impact of using ensemble models on the subchallenge scores. Teams using ensemble models (n=8) vs not using ensemble models (n=5) were compared. We used the Kruskal-Wallis (KW) test for all the comparison. The p value < 0.05 is considered significant.

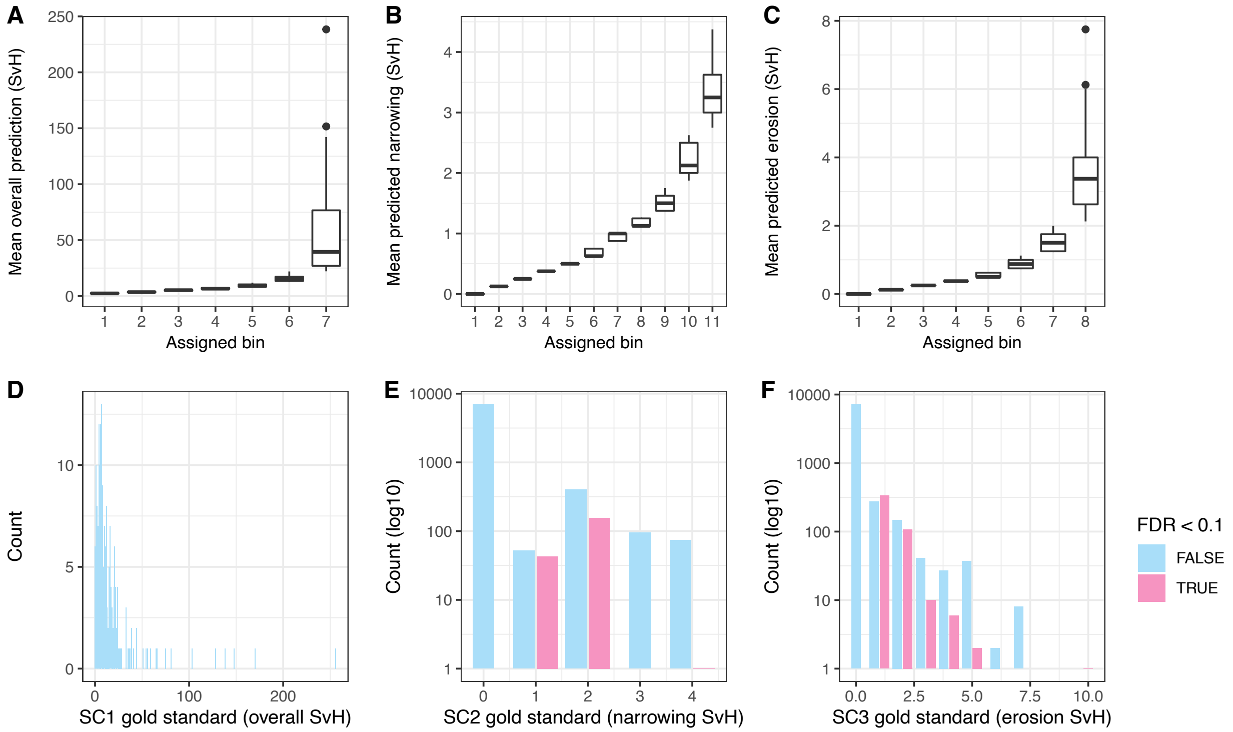


**Figure S7. Identification and correction of gold standard outliers. A-C.** Averaged predictions from top methods were used to assign each measurement to a bin and identify potential outliers (see supplemental methods) in the gold standard dataset for SC1, 2 and 3. **D-F.** False discovery rate (FDR)-adjusted empirical p-values were calculated for each gold standard measurement by comparing them to the rest of the bin they were assigned to. We did not identify any potential outliers in SC1 (overall SvH), but identified several in SC2/SC3 measurements. The potential outlier measurements and the images were reviewed by an expert and, if necessary, corrected (See **Table S4**).

### Supplemental Tables

**Table S1. Links to writeups describing the methods used for all algorithms submitted in the final round.**

**Table S2. Technical questionnaire and responses from the finalist teams.**

**Table S3. Confirmed outlier joints flagged were reviewed and reevaluated by radiology expert in the test set.** The Patient ID (Patient_ID) and joint ID (joint) as well as the original SvH score (score), the revised score after manual review by an expert (score_revised), and the subchallenge the measurement was associated with.

**Table S4. SvH score weights for each patient.** IDs for all patients in the training set as well as the leaderboard and final round test sets (Patient_ID), their overall SvH score (Overall_SvH), and the assigned weight used for that patient in the weighted RMSE.
